# FACS-based detection of extracellular ASC specks from NLRP3 inflammasomes in inflammatory diseases

**DOI:** 10.1101/2024.08.25.24312379

**Authors:** Joanne Topping, Samuel Lara-Reyna, Alice Ibbotson, Heledd Jarosz-Griffiths, Leon Chang, James Poulter, Daniel Peckham, Michael F. McDermott, Sinisa Savic, ImmunAID Consortium

## Abstract

The apoptosis-associated speck-like protein containing a caspase recruitment domain (ASC) is crucial for inflammasome assembly and activation of several inflammasomes, including NLRP3. ASC aggregates are detected in human sera post pyroptotic cell death, but their inflammasome origin remains unclear. This study aimed to develop a method to detect ASC aggregates originating from NLRP3 inflammasomes. Initially, human monocytes, macrophages, and THP-1 ASC reporter cells were employed to validate the detection of ASC/NLRP3-positive events through flow cytometry. The presence of ASC/NLRP3 specks was confirmed in cell supernatants from monocytes and macrophages treated with LPS and nigericin or ATP. Flow cytometry analysis identified double-positive specks in patient sera from inflammatory conditions when compared to healthy controls. Elevated ASC/NLRP3 specks were observed in conditions such as CAPS and Schnitzler’s syndrome. We validated FACS as a reliable method for detecting ASC/NLRP3 specks in human sera, with potential diagnostic and monitoring applications in certain systemic autoinflammatory diseases.

## INTRODUCTION

The NLRP3 (NOD-, LRR- and pyrin domain-containing protein 3) inflammasome is a critical intracellular pattern recognition receptor involved in mediating innate immune responses and release of pro-inflammatory cytokines (Broz & Dixit, 2016). Activation of the NLRP3 inflammasome results in formation of a multimolecular complex containing the sensory component NLRP3, the adaptor protein ASC (apoptosis-associated speck-like protein containing a caspase recruitment domain) and caspase-1 (Broz & Dixit, 2016; Cheng *et al*, 2010). Central to the inflammasome activation is re-localisation of the ASC protein from being diffusely distributed within the cell, to compression into a dense speck in the cytosol within minutes of the inflammasome activation (Cheng *et al*., 2010; Hoss *et al*, 2017). This ASC protein aggregate provides a platform for overall inflammasome organisation, caspase-1 recruitment, and activation, and ultimately as a support for caspase-1 mediated cleavage of pro-interleukin-1 beta (IL-1β) and pro-IL-18 into their active forms (Lara-Reyna *et al*, 2022). The catalytically active ASC specks are released into the extracellular space following cytolytic execution of the pyroptosis (Franklin *et al*, 2014). The presence of these extracellular ASC specks has also been detected in the sera of patients with several conditions where the NLRP3 inflammasome activation has been implicated in the disease pathogenesis (Basiorka *et al*, 2016; Franklin *et al*., 2014; Rowczenio *et al*, 2018; Topping *et al*, 2024).

However, ASC is an adaptor protein also found in other inflammasome complexes including NLRP1, NLRC4, pyrin, AIM2 and NLRP10 (Man & Kanneganti, 2016). The methodologies used for detecting the ASC specks so far have relied on detecting the ASC protein only, without investigating whether ASC was in complex with another inflammasome component. Therefore, these methods were only partially informative, since they could not reliably determine the inflammasome origin of such specks. Previous studies have shown the potential for extracellular ASC specks to be used as a novel biomarkers both for diagnosis and/or disease monitoring (Basiorka *et al*, 2018; Wang *et al*, 2022; Wittmann *et al*, 2023). The diagnostic utility of the extracellular ASC specks could potentially be improved if the inflammasome receptor at the initiation of extracellular specks could be accurately identified. To this end, we developed a novel assay which detects ASC/NLRP3 inflammasome events in the sera of patients with inflammatory conditions.

## RESULTS

To investigate the dynamics of ASC speck release into the extracellular fluid and validate detection methodologies, we employed THP-1 cell lines (human leukemia monocytic cell lines) genetically modified to be either ASC-deficient (THP-1-defASC) or to express ASC tagged with GFP (THP-1-ASC-GFP), with the latter allowing visualization using fluorescence microscopy. Inflammasome induction and ASC speck formation were achieved using a well-established protocol, involving a longer priming phase with LPS, followed by a shorter NLRP3 induction phase using ATP or nigericin. As expected, only THP-1-ASC-GFP cells formed a single ASC speck post-stimulation with LPS and ATP or nigericin (Fig 1A), whereas LPS-only stimulation still induced ASC specks to a lesser extent, with most of the ASC diffused in the cytosol (Fig 1A). To corroborate that ASC specks where secreted into the extracellular space, we imaged the cell-free supernatants of THP-1-ASC-GFP and found ASC specks only in cells stimulated with LPS and ATP or nigericin (Fig 1B). We analysed and confirmed the presence of ASC, by immunoblotting, in THP-1-ASC-GFP intracellularly and in the supernatants (Fig 1C). We then used fluorescence-activated cell sorting (FACS) to detect ASC specks in the cell supernatants using a gating restricted to 1 μm in size (Fig EV1A). While the presence of ASC was minimal in all the controls, and in unstimulated cells and in THP-1-defASC cells (Fig EV1B and C), there was a significant increase in ASC events in THP-1-ASC-GFP cells stimulated with LPS and nigericin (Fig 1D and E). Finally, we detected the presence of mature IL-1β, a marker of inflammasome activation and pyroptotic cell death in the supernatant of THP-1-ASC-GFP stimulated with LPS and nigericin (Fig 1F)

**Figure 1.**
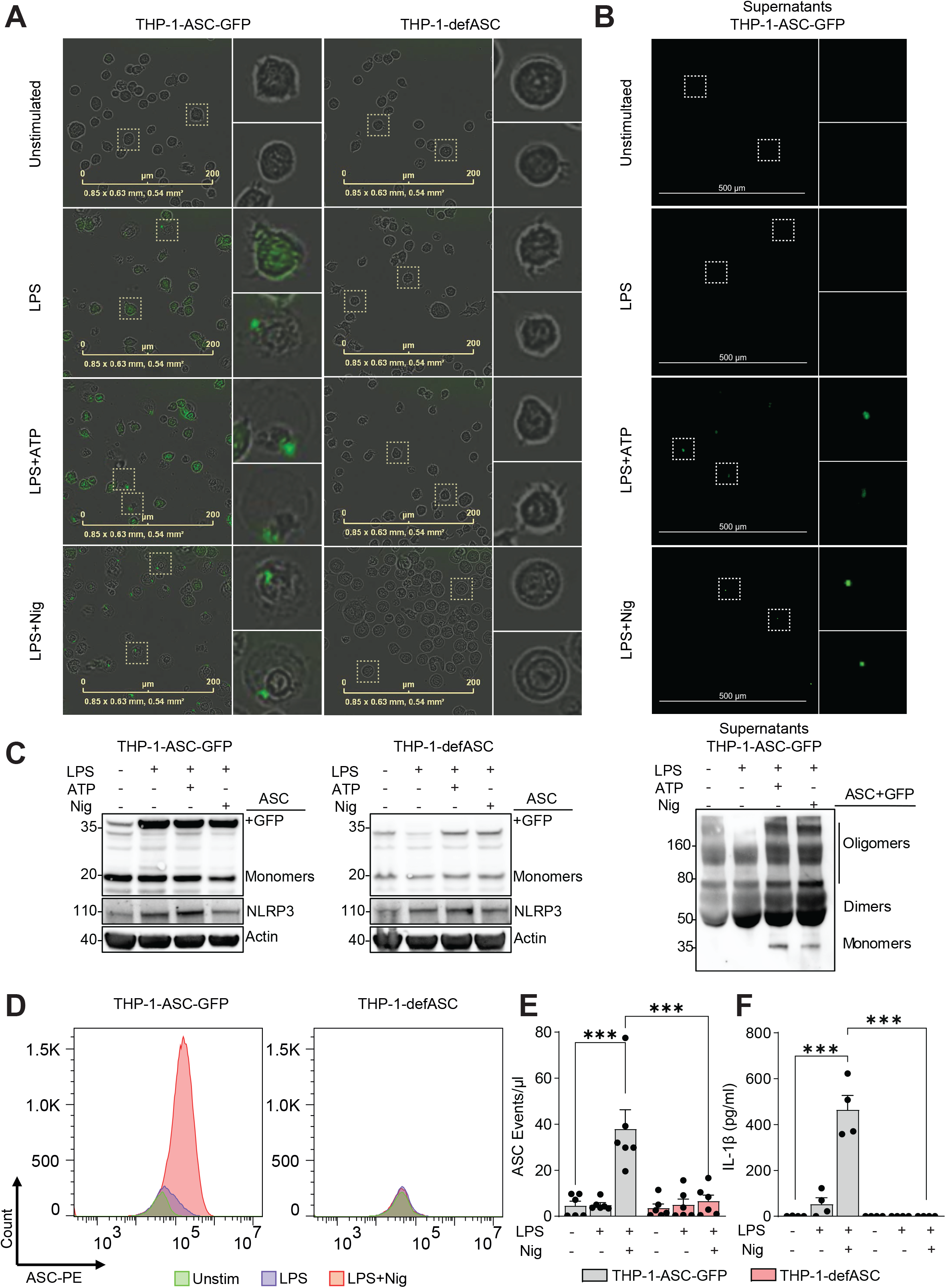
Extracellular ASC in THP-1 cells. **A** Detection of ASC specks in THP-1-defASC or THP-1-ASC-GFP by fluorescent microscopy, stimulated with LPS, LPS + ATP or LPS + nigericin (LPS+ Nig) as indicated in the methods. Each dotted square is shown on the right of the panel as magnified. n=3 independent experiments were carried out. **B**. Extracellular ASC was imaged in the CD7; the images presented here are representative of n=3 independent experiments with 10 images each. **C**. Immunoblots of lysates or supernatants of THP-1-defASC or THP-1-ASC-GFP cells treated as indicated. Images are representative of n = 3 independent experiments. **D** and **E**. ASC specks from THP-1-defASC or THP-1-ASC-GFP cells were captured using flow cytometry, with gating around the 1 μm bead size, showing the ASC events/μL in 30 μL of total volume. F. ELISA assays were used to detect IL-1β in the supernatants of the cells. P-values * =≤ 0.05, ** =≤ 0.01, *** =≤ 0.001 from two-way ANOVA following adjustment for multiple comparisons.

Next, we investigated the co-detection of ASC and NLRP3 proteins in primary human monocytes and macrophages using FACS. Similarly the THP-1 cells, we observed low levels of ASC/NLRP3 specks in all controls and unstimulated monocytes (Fig 2A, and Fig EV2A and B). Stimulation with LPS and ATP or nigericin significantly increased the number of ASC/NLRP3 positive events in the 1 μm and 2 μm gates, but not in the 0.5 μm gate (Fig 2A and B), confirming the presence of ASC/NLRP3 inflammasome complexes. Additionally, immunoblotting confirmed the presence of ASC and NLRP3 in monocytes (Fig 2C). To further validate the detection of ASC/NLRP3 events, we used siRNAs targeting NLRP3 or a non-targeting control (CTRL) in human monocyte-derived macrophages (MDMs) (Fig 2D and E). ASC/NLRP3 events were confirmed in the supernatants of MDMs stimulated with LPS and ATP or nigericin (Fig 2D). These events significantly decreased in MDMs treated with NLRP3 siRNA (Fig 2D), confirming the specific detection of ASC/NLRP3 events. Additionally, we validated the presence of ASC and NLRP3 by immunoblotting in these cells (Fig 2E). Finally, to corroborate our data, we also detected ASC/NLRP3 positive events in the supernatants of THP-1 derived macrophages through FACS and confirmed the presence of ASC and NLRP3 by immunoblotting (Fig EV2D and E)

**Figure 2.**
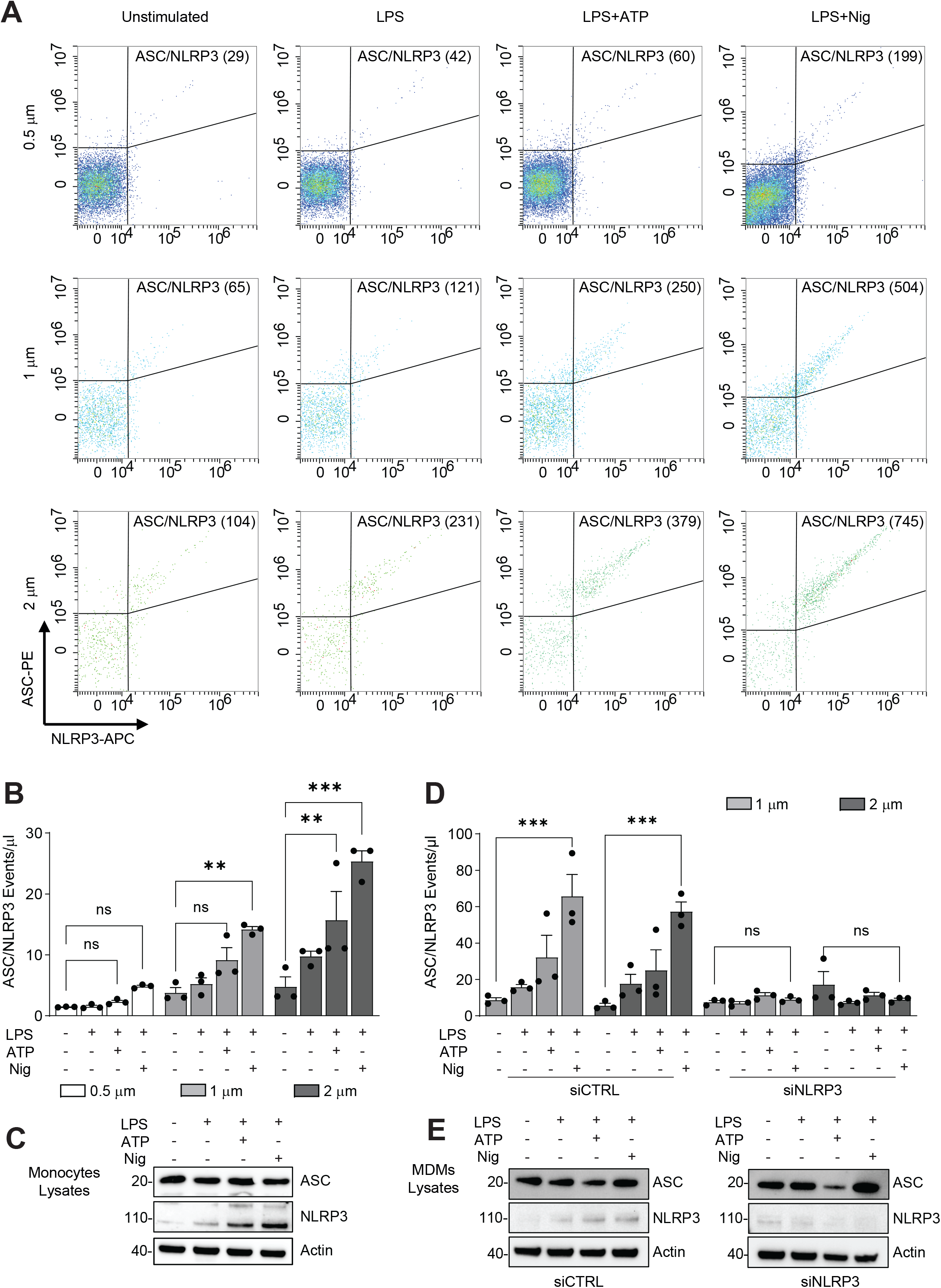
Secretion of ASC/NLRP3 by macrophages. **A and B**. Detection of ASC/NLRP3 specks in the supernatant of primary human monocytes, stimulated as shown in the figure, using flow cytometry with a gating around the 0.5 μm, 1 μm and 2 μm bead size, showing the ASC/NLRP3 events/μL in 30 μL of total volume. **C**. Immunoblots of lysates of primary human monocytes then treated as indicated, immunoblots are representative of n=3 independent experiments. **D**. Detection of ASC/NLRP3 specks in the supernatant of monocyte-derived macrophages (MDMs), stimulated as shown in the figure and transfected with siRNA against NLRP3 or non-targeting control (CTRL). **E**. Immunoblots of lysates of MDMs treated as indicated. Immunoblots are representative of n=3 independent experiments. p-values * =≤ 0.05, ** =≤ 0.01, *** =≤ 0.001 from two-way ANOVA following adjustment for multiple comparisons.

To determine whether ASC specks are detectable under physiological conditions in human sera and if they form complexes with NLRP3, we utilized sera from healthy controls (HC) and patients with inflammatory conditions. Similarly to monocytes and MDMs, serum from systemic autoinflammatory diseases (SAID) showed increased ASC/NLRP3 positive events in the 1 μm and 2 μm gates (Fig 3A and B), but not in the 0.5 μm gate (Fig EV1A), when compared to HC. Interestingly, ASC/NLRP3 positive events were also significantly increased in sera from patients with cystic fibrosis (CF) where events >1 μm were gated (Fig EV3B). We also confirmed the presence ASC and NLRP3 molecules on human sera, with increased levels of these two proteins in SAID and CF patients (Fig EV3C). To assess the diagnostic potential, we tested sera from a large, sex- and age-matched HC population and compared it to conditions associated with NLRP3 inflammasome activation, such as cryopyrin-associated periodic syndrome (CAPS), which is caused by a gain of function mutations in the NLRP3 leading to constitutional NLRP3 inflammasome activity, and Schnitzler’s syndrome, also thought to involve excessive NLRP3 inflammasome activation. Patients with Familial Mediterranean fever (FMF) patients served as controls, given that their condition involves a different inflammasome sensor, pyrin. Our findings revealed that patients with CAPS and Schnitzler’s syndrome exhibit significantly elevated levels of ASC/NLRP3 specks compared to HCs (Fig 3C). Conversely, in FMF patients, ASC/NLRP3 specks were not elevated, aligning with the understanding that the pathogenesis of FMF is more closely associated with a different inflammasome sensor, pyrin (Fig 3C). Finally, to evaluate the reproducibility of ASC/NLRP3 speck detection using this FACS methodology, we analysed 11 individual samples on two separate occasions, more than four weeks apart. Serum sample aliquots were stored at -80°C prior to analyses. The results demonstrated highly consistent levels of ASC/NLRP3 specks across the two time points, indicating that this methodology yields reproducible results and that the analyte remained stable despite undergoing one freeze/thaw cycle and extended storage (Fig 3D).

**Figure 3.**
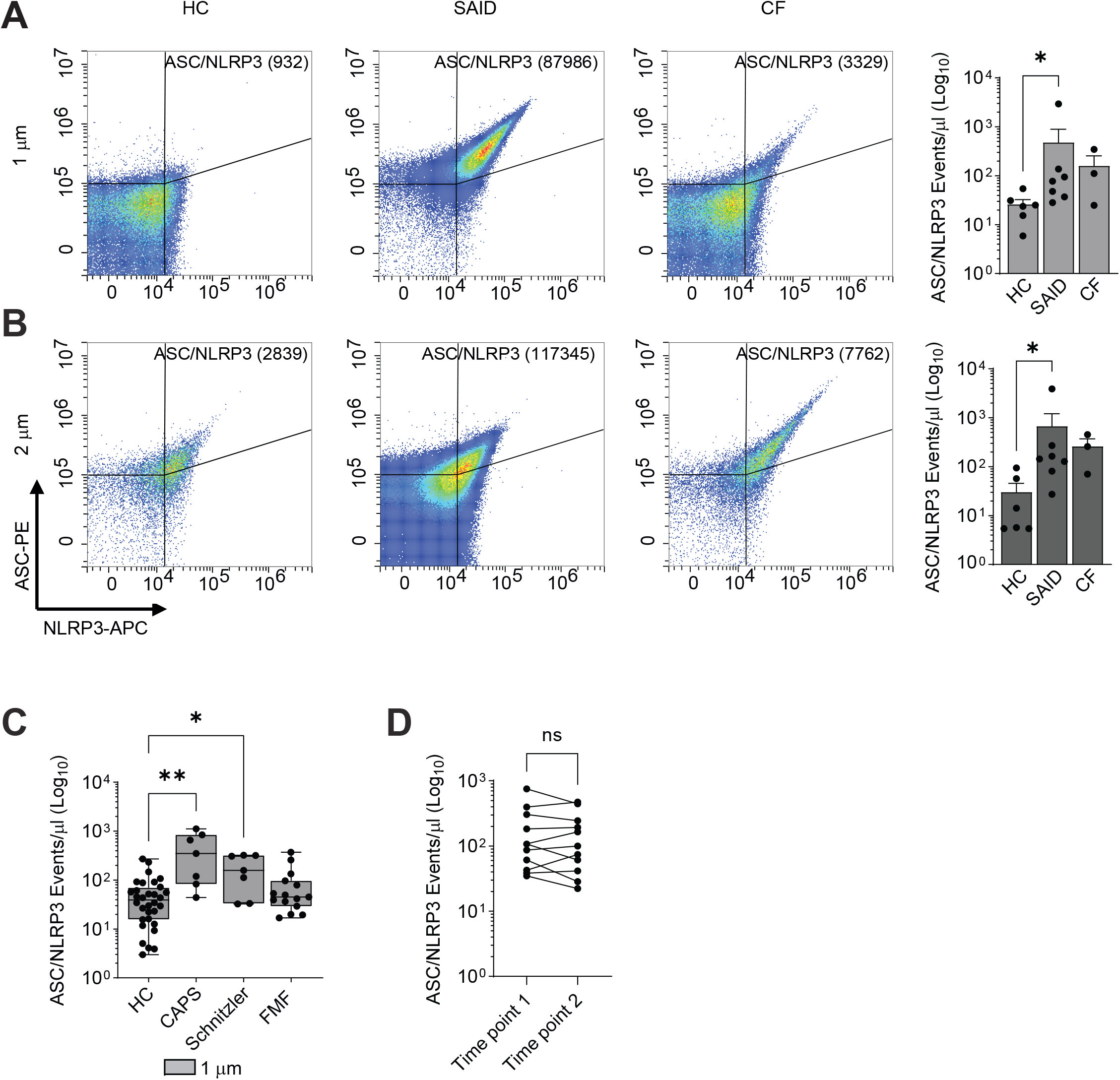
Detection on ASC/NLRP3 in sera of patients with inflammatory disorders. **A and B**. Detection of ASC/NLRP3 specks in the sera of healthy controls (n=6), systemic autoinflammatory diseases (SAIDs) (n=7) and cystic fibrosis (CF) (n=3) patients. Events were recorded using flow cytometry with a gating around the 1 μm and 2 μm bead size, showing the ASC/NLRP3 events/μL in 30 μL of total volume. **C**. Detection of ASC/NLRP3 specks in sera from HC (n=32), CAPS (n=7), Schnitzler’s syndrome (n=7) and Familial Mediterranean Fever (FMF n=15) using FACS based methodology described above. **D**. Independent replicates of 11 serum samples analysed on two dates, more than 4 weeks apart. Samples were aliquoted and frozen at -80°C before analysis. Dots show the mean of duplicate repeats from one frozen aliquot, lines connect the means of the same sample. To look for significant difference between the groups The Kruskal-Wallis test with Dunn’s multiple comparison test was performed, or a Wilcoxon matched-pairs signed rank test was used p-values * =≤ 0.05, ** =≤ 0.01, *** =≤ 0.001.

## DISCUSSION

Inflammasomes are critical intercellular platforms that play essential roles in sensing and propagating signals associated with tissue damage and infection (Lara-Reyna *et al*., 2022). Central to inflammasome activation is the formation of ASC oligomers, which interact with the sensing components defining various inflammasomes (NLRP1, NLRP3, NLRC4, Pyrin, and AIM2), ultimately leading to caspase-1 activation. Caspase-1, in turn, processes several proteins within the cell, including gasdermin D (GSDMD), and the pro-inflammatory cytokines IL-1β and IL-18 (Broz & Dixit, 2016). Several studies have suggested that detecting ASC oligomers (specks) in human sera could serve as a novel method to determine inflammasome activation and act as a useful biomarker. Most of these studies implied that ASC specks originate from NLRP3 inflammasome activation, albeit without providing concrete evidence. To address this gap, we refined our ASC speck detection methodology to investigate their origin.

We developed a FACS-based approach that enables the simultaneous detection of ASC specks in serum and the exploration of their association with other inflammasome components, thereby determining their origin. Initially, we validated our methodology using commercially sourced THP-1 cell lines modified for easy detection of ASC monomers and oligomers by attaching a fluorescent GFP tag to ASC, alongside ASC-deficient cell lines. Using these models, we induced ASC speck formation with traditional inflammasome stimulants and detected specks both intercellularly and in cell culture supernatants using various techniques including IF, WB, and FACS. Extending our experiments to human monocytes and MDMs from healthy controls, we confirmed intercellular ASC specks and their release into cell culture supernatants. Furthermore, we demonstrated the detection of ASC specks in human sera using WB, FACS and visualized these structures with IF. Through WB and FACS, we also identified sensory NLRP3 components within these ASC specks.

This new FACS methodology to detect ASC specks complexed with NLRP3, could facilitate high-throughput analysis suitable for clinical applications. Employing this approach, we observed significantly elevated levels of ASC/NLRP3 specks in patients with constitutively active NLRP3 inflammasomes (CAPS) compared to healthy controls.

In summary, we have used several overlapping methods to develop and validate FACS as a high throughput method for detecting ASC/NLRP3 positive protein specks in human serum. The full diagnostic utility of this assay still needs to be tested in a larger patient cohort and in parallel with other serological markers of inflammation, such as routinely available C-reactive protein (CRP), and inflammatory cytokines, to determine if this biomarker provides any additional diagnostics or prognostic value. We also need to conduct further testing to establish a precise time scale for the degradation of ASC/NLPRP3 specks, which will occur as part of the physiological process. This will help inform the best timing for blood sampling and how the blood samples should be processed and stored. While we have shown that the analyte is stable over a limited period, the conditions for long-term storage conditions still need to be defined. Lastly, using data from the published literature, we developed our FACS method to detect ASC/NLRP3 specks within 1 μm in size. However, it is possible that complexes containing NLRP3, or complexes that have partly degraded, might be outside of this strict size range. Future studies will need to address this point.

## METHODS

### Ethics

Samples were collected from both patients and healthy controls (HC) using Vacuette tubes (Greiner-Bio-One) containing a serum clot activator gel and EDTA for whole blood (50 mL). Patients’ sera were obtained from two distinct sources. Samples were collected from adults with cystic fibrosis (CF) homozygous for F508Δ, known to exhibit elevated levels of serum ASC specks. Ethical approval for the study was granted by the Yorkshire and The Humber Research Ethics Committee (17/YH/0084). Sera from HC and patients with various systemic autoinflammatory conditions, were sourced from the ImmunAID consortium (IMMUNome project consortium for AutoInflammatory Disorders). This study was approved by the South West - Frenchay Research Ethics Committee (REC Reference: 20/SW/0022). Detailed inclusion and exclusion criteria applied for recruiting HC and patients into the ImmunAID project are provided in the supplementary methods.

### Cell lines

THP-1-defASC cells (InvivoGen) or THP-1-ASC-GFP (InvivoGen) were maintained in RPMI with Glutamine (Gibco) supplemented with 10% heat-inactivated FBS (Sigma), and penicillin/streptomycin at 37°C in 5% CO_2_ atmosphere. THP-1 cells were used after rested over-night or differentiated with 50 ng/ml phorbol 12-myristate 13-acetate (PMA, P1585, Sigma) in 24 well plates (Starlab) for 3 days, followed by a 2-day rest period by replacing the differentiation medium with complete medium without PMA. Cells were not used beyond passage 11.

### Human monocytes and MDMs isolation and culture

For isolating primary human monocytes, peripheral blood mononuclear cells (PBMCs) were extracted from EDTA blood tubes using a standard density gradient centrifugation method with Lymphoprep and SepMate tubes (StemCell Tech), as described by the manufacturer. CD14^+^ monocytes were extracted using magnetic microbeads (130-050-201, MACS Miltenyi). Isolated human monocytes were cultured in RPMI with Glutamine (Gibco) supplemented with 10% human AB serum (H4522, Sigma), and penicillin/streptomycin at 37°C in 5% CO2 atmosphere. To differentiate MDMs RPMI medium was supplemented with 20 ng/mL human GM-CSF (PeproTech) for macrophage differentiation and incubated for 6 days, with fresh media replaced on day 3 and day 6. Monocytes were initially seeded at a density of 0.5×10^6^ cells per well in tissue culture-treated 24-well plates (Starlab).

### Cell treatments and siRNAs

Monocytes were allowed to settle overnight prior to experimentation, whereas MDMs were stimulated after differentiation on day 7. Inflammasome activation was achieved by treating the cells with LPS 100 ng/mL (InvivoGen) for 3.5 hours, followed by ATP 5 mM (InvivoGen) or nigericin 20 μM (InvivoGen) for the final 30 minutes of stimulation, for a total of 4 hours. MDMs were transfected with OnTarget Plus smart pool siRNAs for NLRP3 (Dharmacon) or Non-targeting Control Pool (Dharmacon) 24 hours before stimulation, refreshing MDM medium before transfection. All siRNAs were used at a final concentration of 30 nM. The transfection mix was prepared as a 10X mix in OptiMEM containing the appropriate siRNA and TransIT-X2 transfection reagent (MIR 600×, Mirus) in a 1:2 stoichiometry.

### Immunoblotting

For immunoblotting experiments, a total of 0.5×10^6^ cells were seeded into each well of a 24-well plate. Cells were then subjected to differentiation either as MDMs or THP-1 derived macrophages as described above. Upon differentiation, cells were washed with PBS and subsequently lysed on ice for 5 minutes using 80 μl of RIPA buffer supplemented with protease inhibitors (Protease Inhibitor Cocktail set III, EDTA free, Merck) and phosphatase inhibitors (PhosSTOP, Roche). Lysates were then centrifugated at maximum speed for 15 minutes at 4°C, and the cleared lysates were transferred to new tubes. Diluted cleared lysates (1:5) were subjected to the BCA assay (Pierce BCA protein assay kit, 23225, Thermo Scientific) to determine protein concentration. Samples were then diluted in 4X Laemmli sample buffer for use in immunoblotting analyses. Subsequently, equal amounts of protein from each sample were run on 4-12% Bis-Tris gels (Novex, Invitrogen) in MES running buffer and transferred onto PVDF membranes using the iBlot transfer system (Invitrogen). Membranes were blocked with 5% BSA in TBS-T for at least 1 hour at room temperature. Incubation with primary antibodies was carried out overnight at 4°C. Membranes were washed 5 times with TBS-T, then probed with their corresponding secondary antibodies (HRP-Conjugated) diluted at 1:5,000 in 1% BSA in TBS-T, followed by 5 additional washes with TBS-T. Finally, the membranes were incubated with SuperSignal West Pico PLUS chemiluminescent substrate (ThermoFisher) for 1 minute, and the signal was captured using a ChemiDoc Imaging System (BioRad). A list of all antibodies used in this study can be found in Table S1.

### Immunofluorescence cell microscopy

THP-1 cells were recorded after the corresponding stimulations using the IncuCyte Live Cell Analysis system (Sartorius), with the green channel to detect GFP in the THP-1-ASC-GFP. Supernatants were viewed by Structured Illumination Microscopy (SIM) on Zeiss Elyra 7 and Zen Black imaging software.

### Flow Cytometry

For the detection of ASC specks, or protein specks that contained ASC and NLRP3 in a complex, 1.2 μL of PE-conjugated anti-ASC antibody (HASC-71 clone, BioLegend) was used alone, or in combination with 6 μL of APC-conjugated anti-NLRP3 (clone 768319, R&D Systems) added to 60 μL of cell culture media or serum (1:50 and 1:10 dilutions respectively). The mixture was incubated in 96 well plates on a shaker for 1 hour. Size gating was carried out with Flow Cytometry Sub-micron Particle Size beads (ThermoFisher) according to the manufacturer’s specifications and was used to threshold out readings below around 0.5 μm, 1 μm and 2 μm. Samples were run and analysed on a CytoFLEX-S (Beckman Coulter). 30 μL of total volume was acquired in slow settings (10μL/minute).

### Cytokine quantification by ELISA

Cytokines were detected by ELISAs IL-1 beta Human Matched Antibody Pair (ThermoFisher), as per the manufacturer’s recommendations. In general, ELISA plates were coated with 100 μL cytokine capture antibody in PBS overnight at 4°C. The plates were washed three times with PBST (PBS containing 0.5% Tween 20) and the wells were blocked with 300 μL assay buffer (0.5% BSA, 0.1% Tween 20 in PBS) by incubating for 1 hour. The plates were washed twice with PBST, and 100 μL of sera/culture supernatants, together with appropriate standards, were added to wells in duplicates. Immediately, 50 μL of detection antibody was added to all wells and incubated for 2 hours. After incubation, the plates were washed five times with PBST and 100 μL of tetramethylbenzidine (TMB) substrate solution (Sigma) was added to all wells and incubated for 30 minutes. Colour development was stopped by adding 100 μL of 1.8N H_2_SO_4_, and absorbance was measured at 450 nm with a reference at 620 nm. Note that all incubation steps were done at room temperature with continual shaking at 700 rpm. All data points are an average of duplicate technical replicates for each independent experiment.

### Data handling and statistical analysis

Graphs were generated using Prism 11 (GraphPad) and are shown as the mean of N = 3 experiments, each typically consisting of two technical replicates as indicated. Error bars represent the standard error of the mean (SEM). The Kruskal-Wallis test with Dunn’s multiple comparison test was performed when comparing non-parametric populations. A two-way ANOVA statistical test with Tukey’s multiple comparison post-hoc analysis was performed when calculating variance between samples (p values * =≤ 0.05, ** =≤ 0.01, *** =≤ 0.001). Statistical tests used are indicated in the figure legends.

## Supporting information

Methods_inc_exl criteria

ImmunAID investigators

## Data Availability

The data that support the findings of this study are available from the corresponding author, (SS), upon reasonable request.

## AUTHORS CONTRIBUTIONS

SLR, JT and AI conducted experiments. SLR and SS wrote the first draft of the manuscript. SLR, JT, HJZ analysed the data. SS, JP, SLR, JT and MM designed the study. SS, MM and DP secured the funding. All authors read and approved the manuscript.

## Expanded View Figures

**Figure EV1. ASC specks validation on THP-1 cells. A, B and C**. Detection of ASC specks in THP-1-defASC or THP-1-ASC-GFP by FACS, stimulated with LPS, LPS + Nig or using the indicated control, as described in the methods. Validation of ASC specks using flow cytometry with a gating around the 0.5 μm, 1 μm and 2 μm bead size, showing the ASC events/μl in 30 μl of total volume. The number next to the ASC specks represent the total number of events. A total of 6 independent experiments was carried out.

**Figure EV2. ASC/NLRP3 specks on monocytes and THP-1 macrophages. A and B**. Validation of the detection of ASC/NLRP3 specks in human monocytes by FACS; the indicated control is as shown in the figure. Three independent experiments were carried out. Validation of ASC specks using flow cytometry with gating around the 0.5 μm, 1 μm and 2 μm bead size, showing the ASC/NLRP3 events/μl in 30 μl of total volume. The number next to ASC/NLRP3 specks represent the total number of events. **C**. ASC/NLRP3 specks in human monocytes representing the events/μl in 30 μl of total volume combining the events of 1 μm and 2 μm gates. **D**. Detection of ASC/NLRP3 specks in the supernatant of THP-1 macrophages, stimulated as shown in the figure, using flow cytometry with a gating around the 0.5 μm, 1 μm and 2 μm bead size, showing the ASC/NLRP3 events/μl in 30 μl of total volume. **E**. Immunoblots of lysates of THP-1 macrophages then treated as indicated. Immunoblots are representative of n = 3 independent experiments. p values * =≤ 0.05, ** =≤ 0.01, *** =≤ 0.001 from two-way ANOVA following adjustment for multiple comparisons.

**Figure EV3. ASC/NLRP3 specks on patients’ sera. A**. Detection of ASC/NLRP3 specks in the sera of healthy controls (n = 6), systemic autoinflammatory diseases (SAIDs) (n = 7) and cystic fibrosis (CF) (n = 3) patients. Events were recorded using flow cytometry with a gating around the 0.5 μm bead size, showing the ASC/NLRP3 events/μl in a total volume of 30 μl. The number next to ASC/NLRP3 specks represent the total amount of events. **B**. ASC/NLRP3 specks in human sera representing the events/μl in 30 μl of total volume, combining the events of 1 μm and 2 μm gates. **C**. Immunoblots of sera of healthy control, systemic autoinflammatory disease (SAIDs) and cystic fibrosis (CF) patients immunoblots are representative of n = 3 independent experiments. The Kruskal-Wallis test with Dunn’s multiple comparison test was performed, or a Wilcoxon matched-pairs signed rank test. p values * =≤ 0.05, ** =≤ 0.01, *** =≤ 0.001.

