## Supplementary figures and images for "FACS-based detection of extracellular ASC specks from NLRP3 inflammasomes in inflammatory diseases"

### EV Figure 1

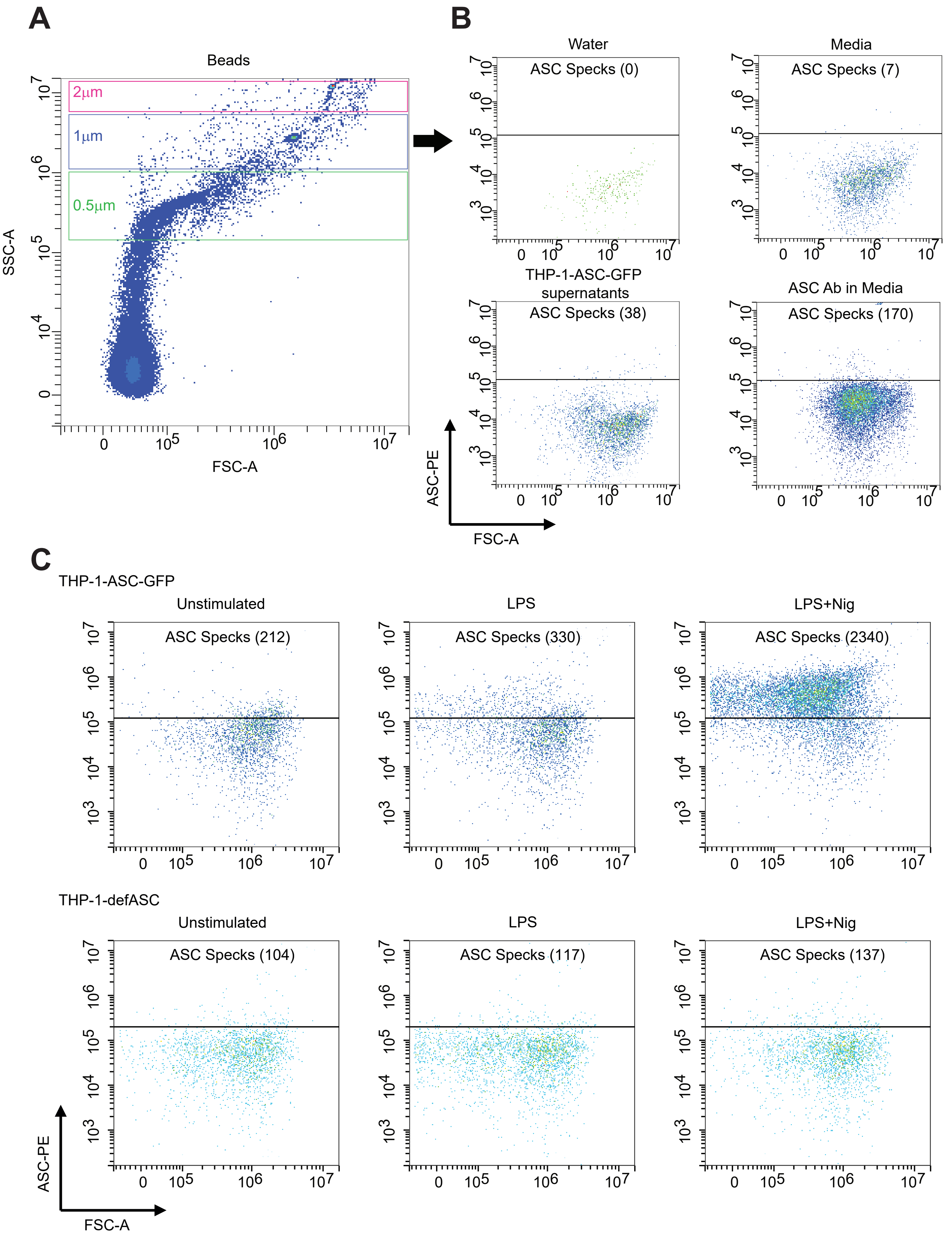


**Extended View (EV) Figure 1**

### EV Figure 2

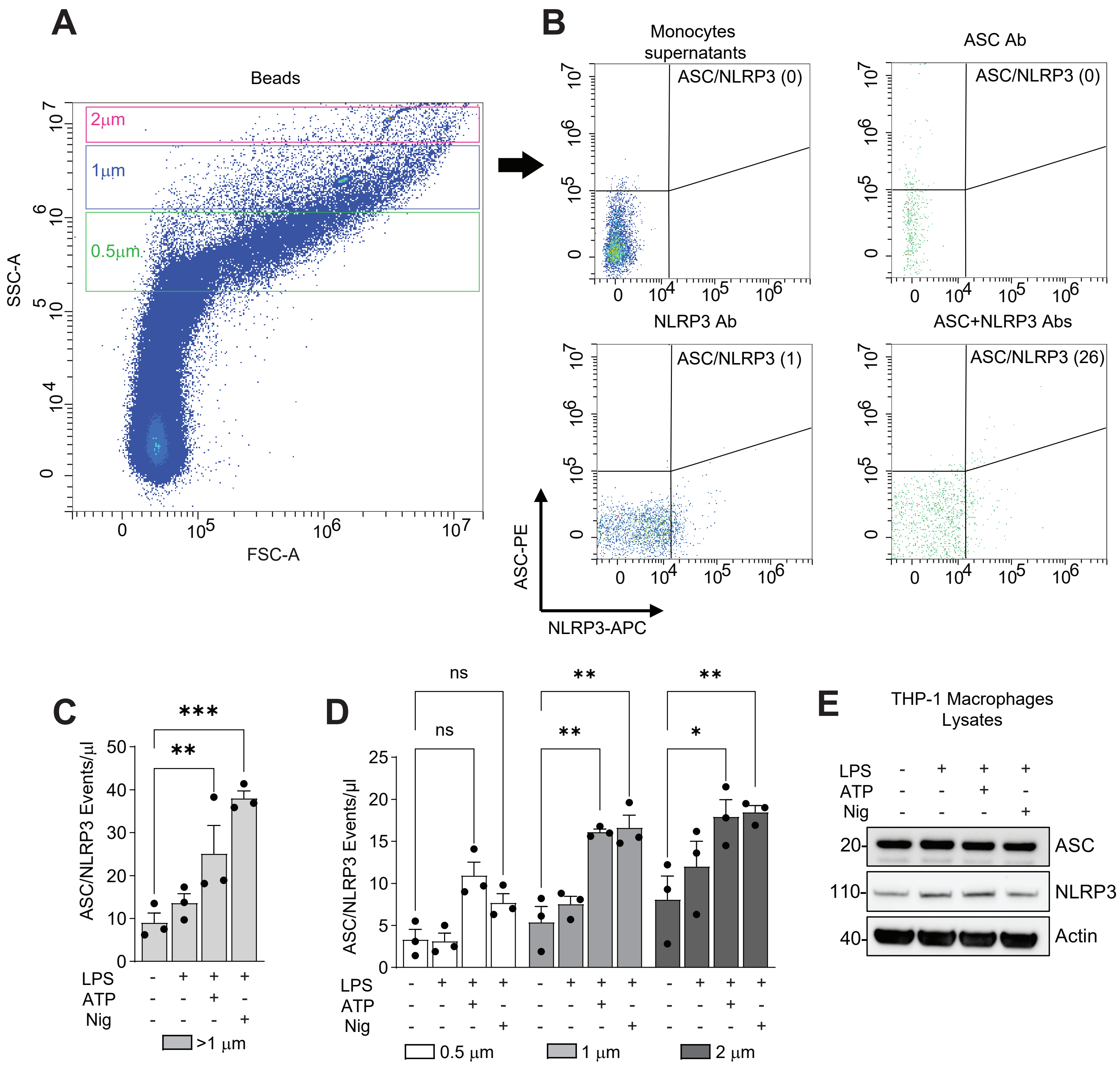


**Extended View (EV) Figure 2**

### EV Figure 3

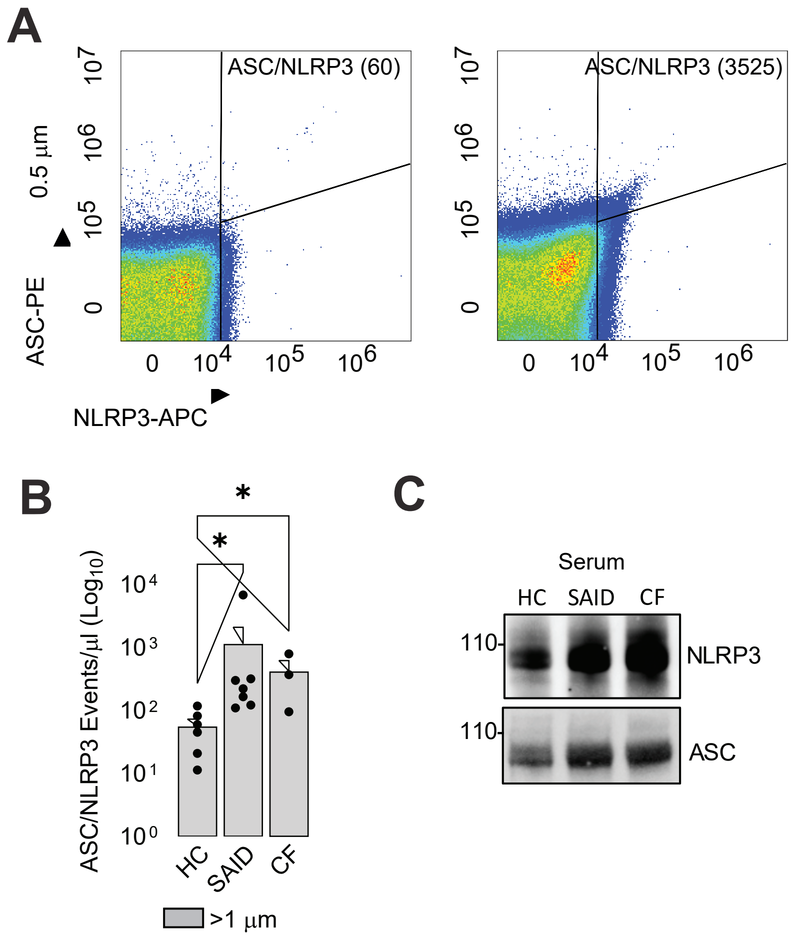


**Extended View (EV) Figure 3**
