## Supplementary material for "FACS-based detection of extracellular ASC specks from NLRP3 inflammasomes in inflammatory diseases": Methods_inc_exl criteria

### ***Inclusion/Exclusion Criteria review***

#### ***Trial : ImmunAID Subgroup 1: Still Diseases : Adult onset Still disease / Systemic Juvenile Idiopathic Arthritis***

This disease can affect people of any age, all patients older than 6 months can be recruited

| Diagnosis Criteria | |
|---|---|
| **Patient ≥16yo**: satisfaction of original Yamaguchi or Fautrel classification criteria | ☐ Yes ☐ No |
| **Patient <16yo**: satisfaction of the ILAR classification criteria for juvenile idiopathic arthritis (2004 release) | ☐ Yes ☐ No |
| **Non-inclusion (must be no)** | |
| Active chronic infection included chronic viral infection (HIV, HBV, HCV…) | ☐ Yes ☐ No |
| Recent infection or antibiotic treatment in the last 2 weeks | ☐ Yes ☐ No |
| Systemic auto-immune disease | ☐ Yes ☐ No |
| Other etiology of fever (infection or neoplasia) | ☐ Yes ☐ No |
| Monogenic auto-inflammatory disease (other than FMF, HIDS, TRAPS, CAPS) | ☐ Yes ☐ No |
| Genetic macrophage activation syndrome | ☐ Yes ☐ No |
| Evidence of immuno-deficiency (e.g., neutropenia, HIV transplant recipient, immunosuppressive treatment for other conditions etc.) | ☐ Yes ☐ No |
| Pregnancy | ☐ Yes ☐ No |
| Individuals deprived of liberty | ☐ Yes ☐ No |
| Inability to understand the local language | ☐ Yes ☐ No |
| Protected persons (under guardianship or curatorship) | ☐ Yes ☐ No |
| **Active disease defined as presence of** | |
| o And CRP ≥ 3 ULN | ☐ Yes ☐ No |
| o And 3 of the following features:<br>▪ arthritis<br>▪ sore throat<br>▪ skin rash<br>▪ lymphadenopathy or splenomegaly<br>▪ increased WBC ≥ 10,000 /mm$^3$<br>▪ PMN ≥ 80%<br>▪ Fever | ☐ Yes ☐ No |
| Date :_____._____._______ | Investigator's Signature |

**Patient number:**______________________ **Patient name:** ______________________

### *Trial : ImmunAID Subgroup 4: Schnitzler*

This disease affects almost exclusively adults

| Diagnosis Criteria | |
|---|---|
| Satisfaction of the Strasbourg classification criteria | ☐ Yes ☐ No |
| **Non-inclusion (must be no)** | |
| Active chronic infection included chronic viral infection (HIV, HBV, HCV…) | ☐ Yes ☐ No |
| Recent infection or antibiotic treatment in the last 2 weeks | ☐ Yes ☐ No |
| Systemic auto-immune disease | ☐ Yes ☐ No |
| Other etiology of fever (infection or neoplasia) | ☐ Yes ☐ No |
| Monogenic auto-inflammatory disease (other than FMF, HIDS, TRAPS, CAPS) | ☐ Yes ☐ No |
| Genetic macrophage activation syndrome | ☐ Yes ☐ No |
| Evidence of immuno-deficiency (e.g., neutropenia, HIV transplant recipient, immunosuppressive treatment for other conditions etc.) | ☐ Yes ☐ No |
| Pregnancy | ☐ Yes ☐ No |
| Individuals deprived of liberty | ☐ Yes ☐ No |
| Inability to understand the local language | ☐ Yes ☐ No |
| Protected persons (under guardianship or curatorship) | ☐ Yes ☐ No |
| **Active disease defined as presence of** | |
| • CRP ≥ 3ULN | ☐ Yes ☐ No |
| 3 of the following features:<br>▪ arthritis<br>▪ urticarial skin rash<br>▪ bone pain<br>▪ lymphadenopathy<br>▪ increased WBC ≥ 10,000 /mm$^3$<br>▪ PMN ≥ 80%<br>▪ Fever | ☐ Yes ☐ No |
| Date :_____._____._______ | Investigator's Signature |

**Patient number:______________________ Patient name: _____________________**

#### *Trial : ImmunAID: mSAID patients*

| Inclusion (must be yes) | |
|---|---|
| Patients with monogenic hereditary SAID: FMF/TRAPS/HIDS/CAPS | ☐ Yes ☐ No |
| Patients diagnosed according to the specific diagnostic criteria of each disease. EUROFEVER criteria *(J Rheumatol 2018, in press)* for clinical definition plus genetic criterion<br>o FMF: Two pathogenic mutations or at least one pathogenic in the exon 10 of the *MEFV* gene<br>o CAPS: One pathogenic mutation in *NLRP3* (or mosaicism)<br>o TRAPS: One pathogenic mutation in *TNFRSF1A* gene<br>o MKD: Two pathogenic mutations in the *MVK* gene or elevated mevalonaturia | ☐ Yes ☐ No |
| 1. Patients with active disease (presence of a flare and/ or persistent chronic inflammation) : ≥ 3 of the 4 following symptoms :<br>o Fever<br>o PhGA ≥ 2/10<br>o Patient AIDAI score ≥ 9 on an at list 29 days period<br>o CRP ≥ ULN (or ≥ 10 mg/L) or /and SAA≥10 mg/L | ☐ Yes ☐ No |
| This disease can affect people of any age, so all patients older than 6 months can be recruited | ☐ Yes ☐ No |
| Patients with health insurance | ☐ Yes ☐ No |
| Signature of the informed consent form (parents/legal representative if the patient is less than <18 years old) | ☐ Yes ☐ No |
| **Non-inclusion (must be no)** | |
| Active chronic infection included chronic viral infection (HIV, HBV, HCV…) | ☐ Yes ☐ No |
| Recent infection or antibiotic treatment in the last 2 weeks | ☐ Yes ☐ No |
| Systemic auto-immune disease | ☐ Yes ☐ No |
| Other etiology of fever (infection or neoplasia) | ☐ Yes ☐ No |
| Monogenic auto-inflammatory disease (other than FMF, HIDS, TRAPS, CAPS) | ☐ Yes ☐ No |
| Genetic macrophage activation syndrome | ☐ Yes ☐ No |
| Evidence of immuno-deficiency (e.g. neutropenia, HIV, transplant recipient, immunosuppressive treatment for other conditions etc.) | ☐ Yes ☐ No |
| Pregnancy | ☐ Yes ☐ No |
| Individuals deprived of liberty | ☐ Yes ☐ No |
| Inability to understand the local language | ☐ Yes ☐ No |
| Protected persons (under guardianship or curatorship) | ☐ Yes ☐ No |
| Date :_____._____.________ | Investigator's Signature |

#### *Trial : ImmunAID: Negative Control*

| Inclusion (must be yes) | |
|---|---|
| Subject free of inflammatory disorders and negative CRP at enrollment | ☐ Yes ☐ No |
| Subject without personal or familial history of SAID | ☐ Yes ☐ No |
| Subject aged from 10 to 60 years old | ☐ Yes ☐ No |
| Subject with health insurance | ☐ Yes ☐ No |
| Signature of the informed consent form (parents/legal representative (if the patient is less than <18 years old) | ☐ Yes ☐ No |
| **Non-inclusion (must be no)** | |
| Active bacterial, viral, fungal, or opportunistic infections | ☐ Yes ☐ No |
| Recent infection or antibiotic treatment in the last 2 weeks | ☐ Yes ☐ No |
| History of any inflammatory, auto-inflammatory or auto-immune disease | ☐ Yes ☐ No |
| History of systemic corticosteroid or non-steroidal (NSAID) therapy within the last 4 weeks | ☐ Yes ☐ No |
| History of neoplasia with the exception of adequately treated basal and squamouscell carcinoma of the skin, or carcinoma in situ of the cervix | ☐ Yes ☐ No |
| Evidence of immunocompromised (e.g. neutropenia, HIV, transplant recipient, immunosuppressive treatment for other conditions etc) | ☐ Yes ☐ No |
| End stage renal disease (eGFR <20 mL/min/1.73m$^2$) | ☐ Yes ☐ No |
| Comorbidities requiring corticosteroid therapy, including those which have required three or more courses of systemic corticosteroids within the previous 12 months | ☐ Yes ☐ No |
| Pregnancy | ☐ Yes ☐ No |
| Current substance abuse or history of substance abuse within the past year. | ☐ Yes ☐ No |
| Lack of peripheral venous access | ☐ Yes ☐ No |
| Individuals deprived of liberty | ☐ Yes ☐ No |
| Inability to understand the local language | ☐ Yes ☐ No |
| Protected persons (under guardianship or curatorship) | ☐ Yes ☐ No |
| Date :_____._____._______ | Investigator's Signature |

**Patient number:______________________ Patient name: _____________________**
