## Supplementary material for "FACS-based detection of extracellular ASC specks from NLRP3 inflammasomes in inflammatory diseases": ImmunAID investigators

| | | |
|---|---|---|
| Vassili Soumelis | | Inserm, U976, Hôpital Saint Louis, Paris, France |
| Arturo Hernandez Cervantes | | Inserm, U976, Hôpital Saint Louis, Paris, France |
| Thomas Henry | | International Center of Infectiology Research (CIRI), University of Lyon, INSERM U1111, Claude Bernard University, Lyon 1, CNRS, UMR5308, ENS of Lyon, Lyon, France |
| Serge Amselem | | Sorbonne Université, INSERM, UMR_S 933, Assistance Publique Hôpitaux de Paris, Département de Génétique médicale, Hôpital Trousseau, Paris, F-75012, France.. |
| Gilles Hayem | | Rheumatology Department. Groupe Hospitalier Paris Saint-Joseph, Paris, France |
| Alexandre Belot | | International Center of Infectiology Research (CIRI), University of Lyon, INSERM U1111, Claude Bernard University, Lyon 1, CNRS, UMR5308, ENS of Lyon, Lyon, France |
| Yvan Jamilloux | | Department of Internal Medicine, University Hospital Croix-Rousse, Hospices Civils de Lyon, University of Lyon 1, Lyon, France; CIRI, Inserm U1111, ENS/CNRS UMR5308, Lyon, France ; Lyon Immunopathology FEderation (LIFE), Lyon, France; Centre de Référence des Maladies Auto-Inflammatoires et de l'Amylose inflammatoire (CEREMAIA), Lyon, France |
| Eric Hachulla | | Univ. Lille, CHU Lille, Département de Médecine Interne et Immunologie Clinique, Centre de Référence des Maladies Systémiques et Auto-Immunes Rares du Nord-Ouest (CERAINO), LIRIC, INSERM, Lille, France |
| Thierry Martin | | Strasbourg University Hospital, Clinical Immunogy department, RESO reference center, Strasbourg France |
| Marie-Elise Truchetet | | Rheumatology Department, Bordeaux University Hospital and Bordeaux University, Bordeaux, France |
| Antoine Néel | | Nantes Université, Internal Medicine Department CHU Nantes, INSERM, Center for Research in Transplantation and Translational Immunology, UMR 1064, F-44000 Nantes, France |
| Bruno Fautrel | | Sorbonne Universités - AP-HP, Groupe Hospitalier Pitié-Salpêtrière, Department of Rheumatology, Centre de Référence des Maladies Auto-Inflammatoires et de l'Amylose inflammatoire (CEREMAIA), F-75013, Paris, France ; INSERM, UMR_S 1136, F-75013 ; CRI-IMIDIATE clinical research network. |
| Nicolas Rosine | nicolas.rosine@aphp..fr | Sorbonne Universités - AP-HP, Groupe Hospitalier Pitié-Salpêtrière, Department of Rheumatology, Centre de Référence des Maladies Auto-Inflammatoires et de l'Amylose inflammatoire (CEREMAIA), F-75013, Paris, France ; INSERM, UMR_S 1136, F-75013 ; CRI-IMIDIATE clinical research network. |
| Emna Chabaane | | Departement Recherche Clinique et Innovation, Hôpital de La Pitiè Salpetrière, Paris, France |
| David Saadoun | | Sorbonne Universités,AP-HP, Groupe Hospitalier Pitié-Salpêtrière, Department of Internal Medicine and Clinical Immunology, Centre de Référence des Maladies Auto-Inflammatoires et de l'Amylose inflammatoire (CEREMAIA), F-75013, Paris, France ; INSERM, UMR_S 959, F-75013; CIC Biotherapie. |
| Matheus Vieira | | Sorbonne Universités,AP-HP, Groupe Hospitalier Pitié-Salpêtrière, Department of Internal Medicine and Clinical Immunology, Centre de Référence des Maladies Auto-Inflammatoires et de l'Amylose inflammatoire (CEREMAIA), F-75013, Paris, France ; INSERM, UMR_S 959, F-75013; CIC Biotherapie. |

| | | |
|---|---|---|
| Isabelle Koné-Paut | | Pediatric rheumatology department and CEREMAIA, Bicêtre hospital APHP, University of Paris Saclay, France |
| Nasima Matsa | | Pediatric rheumatology department and CEREMAIA, Bicêtre hospital APHP, France |
| Perrine Dusser | | Pediatric rheumatology department and CEREMAIA, Bicêtre hospital APHP, University of Paris Saclay, France |
| Jean-David Bouaziz | | APHP, Saint Louis Hospital, Paris Cité University, Paris, France |
| Thibault Mahevas | | APHP, Saint Louis Hospital, Paris Cité University, Paris, France |
| Sophie Georgin-Lavialle | | Sorbonne University, Internal medicine department, Tenon hospital, CEREMAIA, AP-HP, Paris, France |
| Keshia Koutekissa | | Sorbonne University, Internal medicine department, Tenon hospital, CEREMAIA, AP-HP, Paris, France |
| Léa Savey | | Sorbonne University, Internal medicine department, Tenon hospital, CEREMAIA, AP-HP, Paris, France |
| Pierre Quartier | | Université Paris-cité, Paris, France; RAISE reference center for rare diseases, pediatric immunology-hematology and rheumatology unit, Necker hospital, AP-HP, Paris, France |
| Savvas Savvides | | VIB Center for Inflammation Research (IRC), Ghent, Belgium |
| Stephanie Humblet-Baron | | Laboratory of Adaptive Immunology, Department of Microbiology, Immunology and Transplantation, KU Leuven, Leuven, Belgium |
| Adrian Liston | | Department of Pathology, University of Cambridge, Cambridge, United Kingdom |
| Jeroen Raes | | (1) Laboratory of Molecular Bacteriology, Department of Microbiology, Immunology and Transplantation, Rega institute, KU Leuven, Leuven, Belgium (2) VIB Center for Microbiology, Leuven, Belgium |
| Carine Wouters | | Department of Pediatrics, University Hospitals Leuven, Leuven, Belgium; KU Leuven, Department of Microbiology, Immunology and Transplantation, Laboratory of Adaptive Immunology & Immunobiology, Leuven, Belgium |
| Steven Vanderschueren | | Department of General Internal Medicine, University Hospitals Leuven, Leuven, Belgium; KU Leuven, Department of Microbiology, Immunology, and Transplantation, Laboratory of Clinical Infectious and Inflammatory Disorders, Leuven, Belgium |
| Paul Proost | | Laboratory of Molecular Immunology, Department of Microbiology, Immunology and Transplantation, Rega Institute, KU Leuven, Leuven, Belgium |
| Mieke Gouwy | | Laboratory of Molecular Immunology, Department of Microbiology, Immunology and Transplantation, Rega Institute, KU Leuven, Leuven, Belgium |
| Patrick Matthys | | Laboratory of Immunobiology, Department of Microbiology, Immunology and Transplantation, Rega Institute, KU Leuven, Leuven, Belgium |
| Søren Brunak | | Novo Nordisk Foundation Center for Protein Research, University of Copenhagen, Copenhagen N, Denmark |
| Isabella Friis Jørgensen | | Novo Nordisk Foundation Center for Protein Research, University of Copenhagen, Copenhagen N, Denmark |
| Andres Jimenez Kaufmann | | Novo Nordisk Foundation Center for Protein Research, University of Copenhagen, Copenhagen N, Denmark |

| | | |
|---|---|---|
| Sara Garcia | | Novo Nordisk Foundation Center for Protein Research, University of Copenhagen, Copenhagen N, Denmark |
| Philippe Hupé | | Institut Curie, U900 Inserm, PSL Research University, UMR144 CNRS, Paris, France & Mines Paris Tech, Fontainebleau, France |
| Fanny Coffin | | Institut Curie, U900 Inserm, PSL Research University, Paris, France & Mines Paris Tech, Fontainebleau, France |
| Apolline Gallois | | Institut Curie, U900 Inserm, PSL Research University, Paris, France & Mines Paris Tech, Fontainebleau, France |
| Henri de Soyres | | Institut Curie, U900 Inserm, PSL Research University, Paris, France & Mines Paris Tech, Fontainebleau, France |
| Julien Roméjon | | Institut Curie, U900 Inserm, PSL Research University, Paris, France & Mines Paris Tech, Fontainebleau, France |
| Aura Moreno Vega | | OWKIN, Biotec, Paris, France |
| Marc Dubourdeau | | AMBIOTIS SAS - Toulouse - France |
| Karoline Krause | | Charité - Universitätsmedizin Berlin, Institut für Allergieforschung, 12203 Berlin, Germany |
| Dirk Foell | | Department of Pediatric Rheumatology and Immunology, University of Muenster, Muenster, Germany |
| Christoph Kessel | | Department of Pediatric Rheumatology and Immunology, University of Muenster, Muenster, Germany |
| Katerina Laskari | | Rheumatology Unit, 1st Department of Propaedeutic Internal Medicine, Joint Academic Rheumatology Program, University of Athens, Medical School, Athens, Greece |
| Andreakos Evangelos | | Laboratory of Immunobiology, Center for Clinical, Experimental Surgery and Translational Research, Biomedical Research Foundation of the Academy of Athens, Athens, Greece |
| Paul Van Daele | | Department of internal medicine and department of immunology, Erasmus MC, Rotterdam, The Netherlands |
| Clementien Vermont | | Department of internal medicine and department of immunology, Erasmus MC, Rotterdam, The Netherlands |
| Rogier van Wijck | | Department of Pathology & Clinical Bioinformatics |
| Sigrid Swagemakers | | Department of Pathology & Clinical Bioinformatics |
| Peter van der Spek | | Department of Pathology & Clinical Bioinformatics |
| Stefan Erkeland | | Department of Immunology, Erasmus University Medical Center, Rotterdam, The Netherlands |
| Harmen van de Werken | | Department of Immunology, Erasmus University Medical Center, Rotterdam, The Netherlands |
| Yvonne Mueller | | Department of Immunology, Erasmus University Medical Center, Rotterdam, The Netherlands |
| Peter Katsikis | | Department of Immunology, Erasmus University Medical Center, Rotterdam, The Netherlands |
| Michael Hofer | | Pediatric immuno-rheumatology, Dept Pediatrics, CHUV, University Hospital of Lausanne, Lausanne, Switzerland |

| | | |
|---|---|---|
| Cem Gabay | | Division of Rheumatology, Department of Medicine & Department of Pathology and Immunology, HUG and University of Geneva Faculty of Medicine, Switzerland |
| Helen Lachmann | | National Amyloidosis Centre, Royal Free London NHS Foundation Trust and University College London, London, UK |
| Costas Papaloukas | | Department of Biological Applications and Technology, University of Ioannina, Greece |
| Dimitrios I. Fotiadis | | Unit of Medical Technology and Intelligent Information Systems, Department of Materials Science and Engineering, University of Ioannina, Greece |
| Dominique de Seny | | Laboratory of Rheumatology, GIGA-Research, CHU Liège, University of Liège, 4000 Liège, Belgium |
| Haner Direskeneli | | Department of Internal Medicine, Division of Rheumatology, Marmara University, School of Medicine, Istanbul, Turkey |
| Seza Ozen | | Department of Pediatrics, Division of Rheumatology, Hacettepe University Faculty of Medicine, Ankara, Turkey |
| Umut Kalyoncu | | Division of Rheumatology, Department of Internal Medicine, Hacettepe University Faculty of Medicine, Ankara, Turkey |
| Tadej Avcin | | Department of Allergology, Rheumatology and Clinical Immunology, University Children's Hospital, University Medical Centre Ljubljana, Ljubljana, Slovenia |
| Jordi Anton | | Pediatric rheumatology. Hospital Sant Joan de Déu. Institut de Recerca Sant Joan de Déu. Universitat de Barcelona, Spain |
| Violeta Bittermann | | Pediatric rheumatology. Hospital Sant Joan de Déu. Institut de Recerca Sant Joan de Déu. Universitat de Barcelona, Spain |
| Alina Boteanu | | Servicio de Reumatología, Hospital Universitario Ramón y Cajal, Madrid, Spain |
| Fabrizio De Benedetti | | Division of Rheumatology, Bambino Gesù Children's Hospital, IRCCS, Roma, Italy. |
| Antonella Insalaco | | Division of Rheumatology, Bambino Gesù Children's Hospital, IRCCS, Roma, Italy. |
| Emanuele Bizzi | | Internal Medicine Ospedale Fatebenefratelli, Milano, Italy |
| Sonia Caccia | | Department of biomedical and clinical sciences, University of Milano, Italy |
| Luca Cantarini | | Rheumatology Unit Department of Medical Sciences, Surgery and Neurosciences, University of Siena, Siena, Italy. |
| Paolo Sfriso | | Rheumatology Unit, Department of Medicine, University of Padova, Italy. |
| Jean-François Deleuze | | Centre d'Etude du Polymorphisme Humain, Fondation Jean Dausset, Paris, France |
| Sarhan Yaiche | | European Clinical Research Infrastructure Network |
